# Mitochondrial DNA Copy Number Variation in Asthma Risk, Severity, and Exacerbations

**DOI:** 10.1101/2023.12.05.23299392

**Authors:** Weiling Xu, Yun Soo Hong, Bo Hu, Suzy A. A. Comhair, Allison J. Janocha, Joe G. Zein, Ruoying Chen, Deborah A. Meyers, David T. Mauger, Victor E. Ortega, Eugene R. Bleecker, Mario Castro, Loren C. Denlinger, John V. Fahy, Elliot Israel, Bruce D. Levy, Nizar N. Jarjour, Wendy C. Moore, Sally E. Wenzel, Benjamin Gaston, Chunyu Liu, Dan E. Arking, Serpil C. Erzurum, the National Heart, Lung, and Blood Institute (NHLBI) Severe Asthma Research Program (SARP), TOPMed mtDNA Working Group in NHLBI Trans-Omics for Precision Medicine (TOPMed) Consortium

**Author notes:** Address correspondence to Serpil C. Erzurum, M.D. Cleveland Clinic 9500 Euclid Avenue, NC22 Cleveland, Ohio 44195, USA. Co-senior authors. Drs. Serpil C. Erzurum and Dan E. Arking contributed equally to this article. Our study identifies that mitochondrial function plays a role in the molecular basis of asthma. Higher mtDNA-CN, an indicator of mitochondrial function, is predictive of fewer future exacerbations, suggesting that targeting mitochondria and/or metabolism may be useful in future approaches for treatment of exacerbation-prone asthma. Author Contributions: W.X., Y.S.H, A.J.J, D.E.A., and S.C.E. wrote the manuscript. W.X., Y.S.H., B.H., J.G.Z., R.C., and D.E.A. performed data analysis. S.A.A.C. managed samples and assays. S.A.A.C., J.G.Z., D.A.M., D.T.M., V.E.O., E.R.B., M.C., L.C.D., J.V.F., E.I., B.D.L., N.N.J., W.C.M., S.E.W., and B.G. were members of the Severe Asthmatic Research Program, recruited participants, and collected data. D.E.A. performed mitochondrial DNA copy number estimation in the UK Biobank. C.L. led the TOPMed mtDNA Working Group in NHLBI Trans-Omics for Precision Medicine (TOPMed) and supported mitochondrial DNA copy number estimation of Severe Asthma Research Program. V.E.O. led the TOPMed Asthma Working Group. All authors have read and agreed to the published version of the manuscript. This research was conducted using the UK Biobank Resource under Application Number 17731. The study was supported by the National Heart, Lung, and Blood Institute (HL081064, HL103453, and HL144569) and the National Center for Advancing Translational Sciences (ULTR000439). SARP was supported by awards from the National Heart, Lung, and Blood Institute (U10 HL109172, U10 HL109168, U10 HL109152, U10 HL109257, U10 HL109046, U10 HL109250, U10 HL109164, U10 HL109086). SARP mitochondrial DNA copy numbers were from the Trans-Omics in Precision Medicine (TOPMed) program supported by the National Heart, Lung and Blood Institute (NHLBI). Core support including centralized genomic read mapping and genotype calling, along with variant quality metrics and filtering were provided by the TOPMed Informatics Research Center (3R01HL117626-02S1; contract HHSN268201800002I). Core support including phenotype harmonization, data management, sample-identity quality control, and general program coordination were provided by the TOPMed Data Coordinating Center (R01HL120393; U01HL120393; contract HHSN268201800001I). SARP3 disclosure statement (industry): The following companies provided financial support for study activities at the Coordinating and Clinical Centers beyond the third year of patient follow-up: AstraZeneca, Boehringer-Ingelheim, Genentech, GlaxoSmithKline, Sanofi-Genzyme-Regeneron, and TEVA. These companies had no role in study design or data analysis, and the only restriction on the funds was that they be used to support the SARP initiative.

## Abstract

**Rationale:** Although airway oxidative stress and inflammation are central to asthma pathogenesis, there is limited knowledge of the relationship of asthma risk, severity, or exacerbations to mitochondrial dysfunction, which is pivotal to oxidant generation and inflammation.

**Objectives:** We investigated whether mitochondrial DNA copy number (mtDNA-CN) as a measure of mitochondrial function is associated with asthma diagnosis, severity, oxidative stress, and exacerbations.

**Methods:** We measured mtDNA-CN in blood in two cohorts. In the UK Biobank (UKB), we compared mtDNA-CN in mild and moderate-severe asthmatics to non-asthmatics. In the Severe Asthma Research Program (SARP), we evaluated mtDNA-CN in relation to asthma severity, biomarkers of oxidative stress and inflammation, and exacerbations.

**Measures and Main Results:** In UK Biobank, asthmatics (*n* = 29,768) have lower mtDNA-CN compared to non-asthmatics (*n* = 239,158) (beta, -0.026 [95% CI, -0.038 to -0.014], *P* = 2.46×10^-5^). While lower mtDNA-CN is associated with asthma, mtDNA-CN did not differ by asthma severity in either UKB or SARP. Biomarkers of inflammation show that asthmatics have higher white blood cells (WBC), neutrophils, eosinophils, fraction exhaled nitric oxide (F_E_NO), and lower superoxide dismutase (SOD) than non-asthmatics, confirming greater oxidative stress in asthma. In one year follow-up in SARP, higher mtDNA-CN is associated with reduced risk of three or more exacerbations in the subsequent year (OR 0.352 [95% CI, 0.164 to 0.753], *P* = 0.007).

**Conclusions:** Asthma is characterized by mitochondrial dysfunction. Higher mtDNA-CN identifies an exacerbation-resistant asthma phenotype, suggesting mitochondrial function is important in exacerbation risk.

## Introduction

One of the most common chronic diseases in the United States, asthma is characterized by airway inflammation and hyper-reactivity. Severe disease, which occurs in 10% of asthmatics, is difficult to treat, and those with frequent exacerbations, which occur in 5% of asthmatics, suffer a disabling burden with asthma. Thus, prevention of exacerbations is the major goal of global asthma care guidelines. Yet, we lack understanding of the molecular mechanisms that lead to the exacerbation-prone asthma phenotype and thus, are unable to apply specific prevention interventions. Pre-clinical models and human studies identify excess production of reactive oxygen species, with bursts of oxidants during exacerbations (1-6). Although mitochondria are pivotal to oxidant generation and inflammation, and changes in mitochondria function are linked to asthma (3, 7), the relationship of asthma risk, severity, or exacerbations to mitochondria is unknown.

Mitochondria produce energy from oxidative-phosphorylation and regulate redox status. Each human cell contains hundreds to thousands of mitochondria, dependent on tissue and cell type (8). Mitochondria have their own circular DNA (mtDNA) that contains 13 genes encoding oxidative phosphorylation proteins crucial for bioenergetics, but the majority of oxidative phosphorylation proteins are encoded by nuclear DNA. Mitochondria independently replicate, transcribe, and translate their mtDNA using their own ribosomal and transfer RNA, which are encoded by mtDNA. Thus, reductions in mtDNA-CN lead to low levels of mitochondrial-encoded proteins in oxidative phosphorylation, decreased respiratory capacity, and greater production of reactive oxygen species, all of which lead to immune dysfunction, inflammation, and altered cell signaling (9, 10). Thus, mitochondrial DNA copy number (mtDNA-CN) is a good proxy of mitochondrial function, with low levels indicating mitochondrial dysfunction. Variation of mtDNA copy number (mtDNA-CN) has been successfully used as an indirect assessment of mitochondrial function in population studies (9, 10), i.e., low mtDNA-CN has been associated with cardiovascular disease, diabetes, and cancers (9, 11).

We hypothesize that mitochondrial dysfunction is associated with asthma and the parameters of severity and exacerbation. To test if mitochondrial dysfunction is associated with asthma, we analyzed mtDNA-CN estimated from a combination of whole exome sequencing data and genotyping data in participants of the UK Biobank (UKB). To carefully determine whether lower mtDNA-CN is associated with asthma severity and whether there is a relationship to exacerbations, we analyzed mtDNA-CN, measures of inflammation and oxidative stress, and asthma exacerbations in severe and non-severe asthmatics of the large-scale consortium-based Severe Asthma Research Program (SARP) through NHLBI Trans-Omics for Precision Medicine (TOPMed).

## Methods

### Study Population

The curated UKB dataset includes 268,926 unrelated individuals who self-identify as White (European) ethnicity including 29,768 asthmatics and 239,158 non-asthmatics. Asthmatics were defined based on self-report (12). The UKB curated exclusion criteria are detailed in the supplement and in prior reports [see UKB criteria in supplement] (13). Exclusions specific to these analyses include current smokers, cancer, other lung diseases, gastro-esophageal reflux disease, chronic sinusitis, sleep apnea, and kidney, liver, and heart failure [supplement, Table E1]. All study participants provided written informed consent, and all centers obtained approval from their institutional review boards (13). Normative lung functions were predicted to calculate the % FEV1 in the UKB (supplement, UKB lung function methods, Table E2). Mild or moderate-severe asthma were defined by asthma medication use id codes in the UKB dataset [supplement] (12).

The Severe Asthma Research Program (SARP) includes 1283 asthmatics, including 703 non-severe and 580 severe asthmatics as previously defined [see supplement, SARP dataset] (14, 15). Exclusion criteria similar to those in the UKB are detailed in the supplement. Fractional exhaled nitric oxide (F_E_NO) was measured by an online method at a constant flow rate of 50 ml/second according to the standards published by ATS. Asthma exacerbation was defined as an event of a worsening of asthma requiring treatment with systemic corticosteroids for three or more days to prevent a serious outcome. Asthma exacerbations in the prior year were determined by participant recall for the past twelve months. For exacerbations in the one-year longitudinal follow up, exacerbation events were collected using standardized questionnaires at in-person visits or during phone calls. Those with three or more exacerbations in the one-year follow up have been designated as an exacerbation-prone asthma phenotype (16). The study protocol and procedures were approved by the IRB at each participating center and by an independent Data Safety Monitoring Board. All participants provided written informed consent. The study is registered on ClinicalTrials.gov [NCT01750411].

### Mitochondrial DNA Copy Number (mtDNA-CN)

DNA used for mtDNA-CN estimation in the UKB was derived from buffy coat. mtDNA-CN was estimated from a combination of whole exome sequencing data and probe intensities from the Affymetrix Axiom array (13). DNA used for mtDNA-CN estimation in SARP was derived from whole blood. Samples underwent whole genome sequencing (WGS) at the New York Genome Center of TOPMed (https://topmed.nhlbi.nih.gov/). mtDNA-CN was estimated from WGS by comparing the coverage of mitochondrial reads to autosomal reads using the program fastMitoCalc in mitoCaller (17). Raw unadjusted means of mtDNA-CN are provided for the UKB and SARP and termed mtDNA-CN. Because there is variation in mtDNA-CN methodology (Affymetrix Axiom array in the UKB as opposed to mtDNA/nDNA in TOPMed) and because depending on study, mtDNA-CN and age, sex, and white blood cell (WBC) measurement can vary, we calculated mtDNA-CN residuals after adjusting for covariates that may influence mtDNA-CN and standardized these residuals to a mean of 0 and SD of 1. These include age, sex, center where sample was collected, and WBC as well as neutrophils, eosinophils, and/or platelets when available (Table 1) (see supplement, Adjusted mtDNA-CN).

**Table 1.** Covariates used to adjust mtDNA-CN.

|  | Cohort | Collection Center | Age | Sex | Ethnicity | Covariate<br>White blood cells | Neutrophils | Eosinophils | Platelets | Red Blood Cells |
| --- | --- | --- | --- | --- | --- | --- | --- | --- | --- | --- |
| UKB | NA* | + | + | + | NA** | - | + | + | + | + |
| SARP | + | + | + | + | + | + | + | - | NA*** | - |

(+) indicates covariate is used; (-) indicates covariate not used; NA\*, Not applicable in the UKB which did not have cohorts; NA\*\*, Not applicable for the UKB as only whites were evaluated, so ethnicity is not adjusted; NA\*\*\*, Platelet counts are not available for individuals in SARP. Associations tested include: (1) association between asthma status and mtDNA-CN adjusted for age, age<sup>2</sup>, sex, center, natural log-transformed neutrophil and eosinophil counts, and platelet counts, and (2) the association between asthma status and mtDNA-CN adjusted for non-linear age (restricted cubic splines with 3 knots), asthma status, an interaction between non-linear age and asthma status, sex, center, log-transformed neutrophil and eosinophil counts, and platelet counts. See supplemental methods for details.

### Serum Antioxidant Analyses

Superoxide dismutase (SOD) activity and glutathione peroxidase activity (GPx) were determined as previously described (18, 19).

### Statistical Analysis

Analyses were conducted with JMP Pro 15 (SAS Institute, Cary, NC) or R (version 4.2.2, cran-project.org). Data were summarized as mean ± SD or frequency (percentage) where appropriate. For comparisons of two groups, Student’s t-test or Wilcoxon test was used as appropriate. For three or more groups, analysis of variance (ANOVA) was used. Non-normally distributed variables were log transformed prior to linear regression analysis. The level of significance for *P* was chosen at 0.05.

### Association with Asthma Phenotypes

Models were adjusted as appropriate to address hypotheses (Table 1) (see supplement, Adjusted mtDNA-CN). For the UKB, the differences in mtDNA-CN (95% confidence intervals [CI]) by presence of asthma and by asthma severity were evaluated using linear regression models adjusted for age, age^2^, sex, center, log-transformed neutrophil and eosinophil counts, and platelet count (supplement). To examine the relationship between unadjusted mtDNA-CN levels and age by asthma, models were used to include an interaction term between age and asthma, adjusting the same set of covariates (Table 1 and supplement). To also allow for non-linear association between age and mtDNA-CN, we used age as a continuous variable with restricted cubic splines with 3 knots (at 10^th^, 50^th^, and 90^th^ percentiles). We performed separate analyses for asthma (no / yes) and for asthma severity (non-asthma / mild asthma / moderate-severe asthma). For SARP, the difference (95% CI) in mtDNA-CN by non-severe and severe asthma, and the difference (95% CI) in mtDNA-CN by exacerbation occurrence were examined using linear regression models adjusted for age, age^2^, sex, ethnicity, WBC and neutrophil counts, SARP cohort, and cohort and age interaction (Table 1 and supplement). The relationship between mtDNA-CN and age by asthma severity was examined using linear regression models adjusted for sex, ethnicity, WBC and neutrophil counts, and SARP cohort (Table 1 and supplement).

## Results

### Asthma in UK Biobank

The clinical characteristics of participants in the UKB are displayed in Table 2. The asthma group has a greater percentage of females, lower lung functions, and higher neutrophil and eosinophil counts (*P <* 0.001 for all) (Table 2), consistent with known characteristics of asthma. Based on medication use, among the asthmatics, 25,322 (85%) have mild asthma and 4,446 (15%) have moderate-severe asthma. Compared with mild asthmatics, moderate-severe asthmatics are older, have higher body mass index (BMI), lower lung functions, and higher eosinophil counts (Table 2), consistent with known characteristics of severe asthma. Asthmatics have lower unadjusted mtDNA-CN (Table 2). Even after taking into account covariates of age, sex, and blood cell counts (see Table 2 and supplement), mtDNA-CN is significantly lower in asthmatics compared to non-asthmatics (beta, -0.026 [95% CI, -0.038 to -0.014], *P* = 2.46×10^-5^) (Table 2) (Figure 1). Both mild and moderate-severe asthmatics have lower adjusted mtDNA-CN compared to non-asthmatics (mild asthma, beta, -0.024 [95% CI, -0.037 to -0.012], *P* = 1.90×10^-4^; moderate-severe asthma, beta, -0.034 [95% CI, -0.063 to -0.004], *P* = 0.02) (Figure 1). Moderate-severe asthmatics have lower unadjusted mtDNA-CN than mild asthmatics (Table 2), but there is no significant difference in mtDNA-CN by severity when adjusted for age, age^2^, sex, center, log-transformed neutrophil and eosinophil counts, and platelet counts (Table 2) (Figure 1). mtDNA-CN is inversely related to age in asthmatic and non-asthmatic individuals (all *P <* 0.001). The decline in mtDNA-CN with increasing age is similar among groups (all *P* > 0.05) (Figure 2A – D). Overall, mtDNA-CN is lower in asthmatics as compared to non-asthmatics and is independent of asthma severity defined by medication use. To more carefully assess mitochondrial dysfunction among non-severe and severe asthma, we examined mtDNA-CN SARP network data.

**Figure 1.**
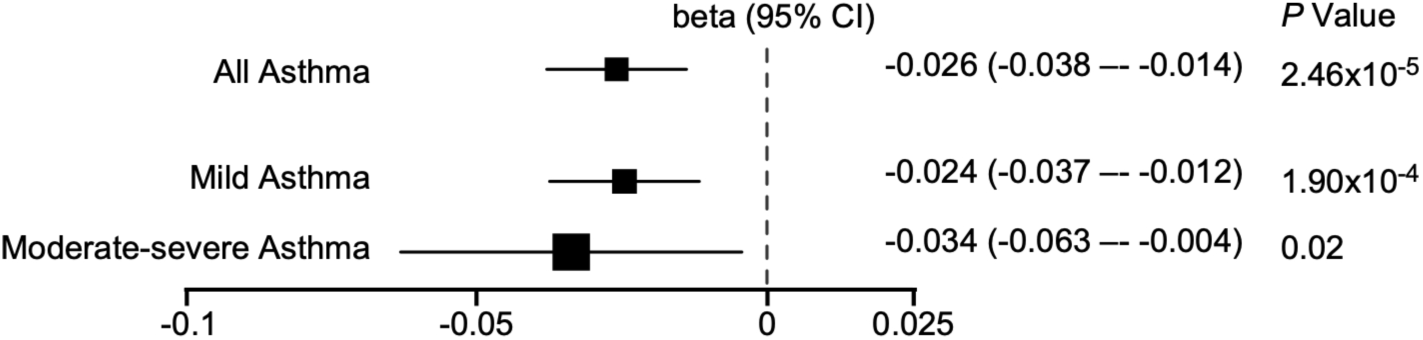
Asthmatics have lower mitochondrial DNA copy number (mtDNA-CN) compared to non-asthmatics in the UK Biobank (UKB). Forest plot depicting beta value and associated 95% confidence interval (CI) of mtDNA-CN in all asthmatics, mild or moderate-severe asthmatics as compared to non-asthmatics.

**Figure 2.**
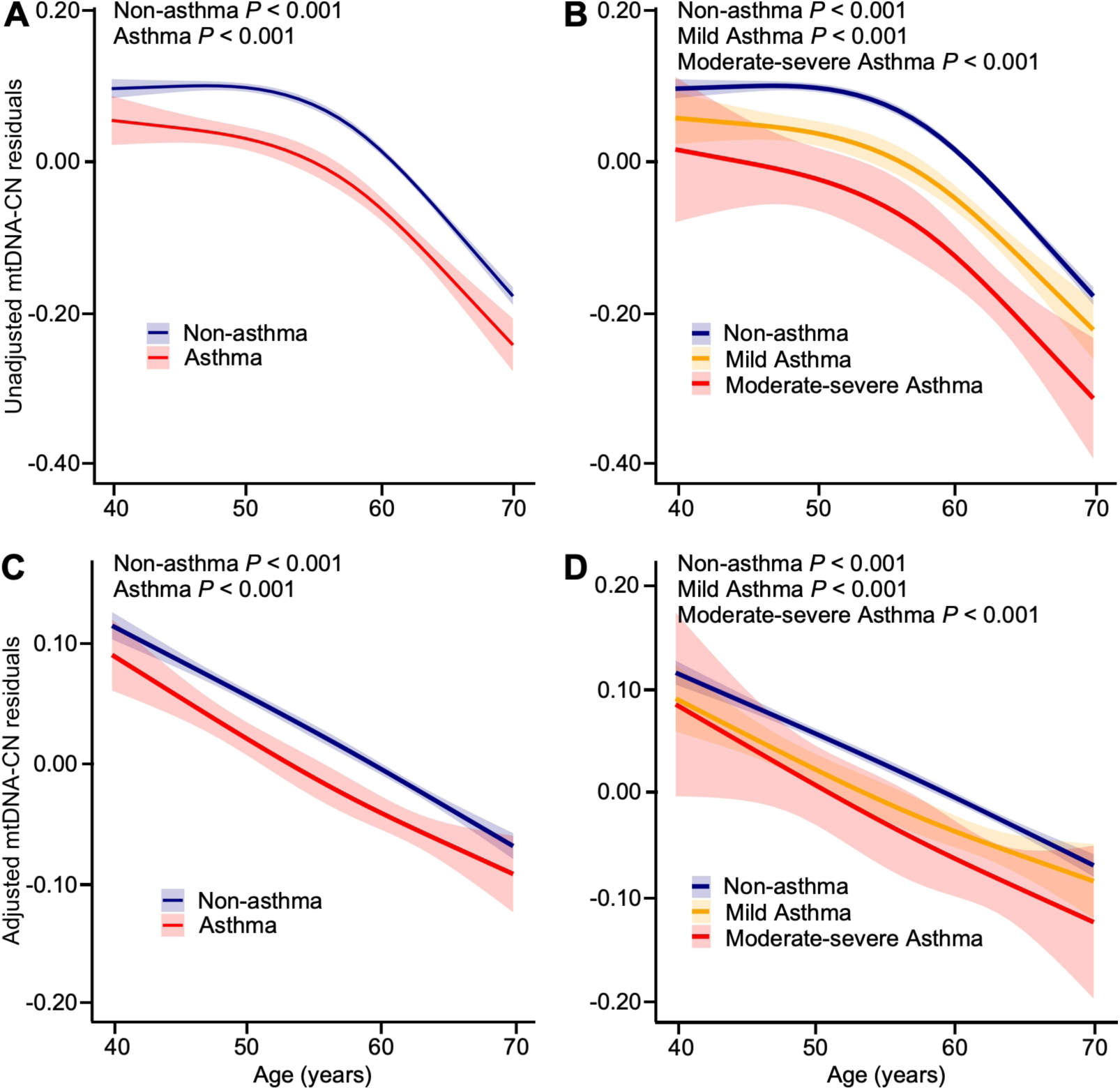
mtDNA-CN inversely related with age in the UKB. (*A-B*) The predicted mtDNA-CN residuals using the unadjusted model (including only non-linear age (restricted cubic splines with 3 knots), asthma status, and an interaction between non-linear age and asthma status) decreases with age in non-asthmatics and all asthmatics (*A*), and in mild asthmatics, and moderate-severe asthmatics (*B*). (*C-D*) The predicted mtDNA-CN residuals marginally adjusted for sex, center, log-transformed neutrophil and eosinophil counts, and platelet counts were significantly related with age in non-asthmatics and all asthmatics (*C*), and in mild asthmatics, and moderate-severe asthmatics (*D*).

**Table 2.** Features of asthmatics and non-asthmatics in the UK Biobank (UKB)

| Characteristics | Non-asthma<br>(n = 239,158) | Asthma<br>(n = 29,768) | P<br>Value* | Mild Asthma<br>(n = 25,322) | Moderate-severe<br>Asthma<br>(n = 4,446) | P<br>Value** | P<br>Value*** |
| --- | --- | --- | --- | --- | --- | --- | --- |
| Age — yr | 56 ± 8 | 55 ± 8 | <0.0001 | 55 ± 8 | 57 ± 8 | <0.0001 | <0.0001 |
| Female — % | 54 | 57 | <0.0001 | 56 | 61 | <0.0001 | <0.0001 |
| Body mass index — kg/m<sup>2</sup> | 27.2 ± 4.6 | 27.9 ± 5.2 | <0.0001 | 27.8 ± 5.0 | 28.6 ± 5.6 | <0.0001 | <0.0001 |
| Blood pressure — mm Hg |  |  |  |  |  |  |  |
| Systolic | 138 ± 18.6 | 137 ± 17.9 | <0.0001 | 137 ± 17.8 | 140 ± 18.2 | <0.0001 | 0.0011 |
| Diastolic | 82 ± 10.1 | 82 ± 9.9 | 0.0006 | 82 ± 9.9 | 83 ± 9.7 | 0.0003 | <0.0001 |
| Hemoglobin — g/dL | 14.2 ± 1.2 | 14.2 ± 1.2 | 0.07 | 14.2 ± 1.2 | 14.1 ± 1.2 | 0.0008 | 0.005 |
| Hematocrit — % | 41.1 ± 3.4 | 41.1 ± 3.4 | 0.9 | 41.1 ± 3.4 | 41.0 ± 3.4 | 0.02 | 0.5 |
| Lung functions<sup>†</sup> |  |  |  |  |  |  |  |
| Forced expiratory volume<br>in 1 second — % predicted | 97.6 ± 16.2 | 89.4 ± 18.3 | <0.0001 | 90.1 ± 18.0 | 84.9 ± 19.6 | <0.0001 | <0.0001 |
| Forced vital capacity —<br>liters | 3.9 ± 1.0 | 3.7 ± 1.0 | <0.0001 | 3.7 ± 1.0 | 3.4 ± 0.9 | <0.0001 | <0.0001 |
| FEV<sub>1</sub>/FVC | 0.77 ± 0.06 | 0.73 ± 0.08 | <0.0001 | 0.74 ± 0.08 | 0.71 ± 0.10 | <0.0001 | <0.0001 |
| White blood cells —<br>x10<sup>9</sup>/liter | 6.7 ± 1.6 | 6.9 ± 1.7 | <0.0001 | 6.9 ± 1.7 | 7.3 ± 1.8 | <0.0001 | <0.0001 |
| Neutrophils | 4.1 ± 1.3 | 4.3 ± 1.4 | <0.0001 | 4.3 ± 1.4 | 4.6 ± 1.5 | <0.0001 | <0.0001 |
| Eosinophils | 0.16 ± 0.12 | 0.22 ± 0.16 | <0.0001 | 0.22 ± 0.15 | 0.25 ± 0.18 | <0.0001 | <0.0001 |
| Platelets — x10<sup>9</sup>/liter | 251 ± 56 | 259 ± 58 | <0.0001 | 257 ± 58 | 269 ± 60 | <0.0001 | <0.0001 |
| mtDNA-CN | 67.8 ± 12.7 | 67.1 ± 12.7 | <0.0001 | 67.3 ± 12.6 | 66.1 ± 12.8 | <0.0001 | <0.0001 |
| Unadjusted mtDNA-CN<br>residuals | 0.020 ± 0.99 | -0.040 ± 0.99 | <0.0001 | -0.02 ± 0.98 | -0.118 ± 1.00 | <0.0001 | <0.0001 |
| Adjusted mtDNA-CN<br>residuals | 0.005 ± 0.99 | -0.021 ± 0.99 | <0.0001 | -0.020 ± 0.99 | -0.030 ± 1.01 | 0.5 | <0.0001 |

Mean ± SD \*P value for non-asthmatics vs. asthmatics. \*\*P value for mild asthmatics vs.
moderate-severe asthmatics. \*\*\*P value for ANOVA of non-asthmatics, mild asthmatics,
and moderate-severe asthmatics. <sup>†</sup>Lung function available in subset where testing was
done. <sup>‡</sup>Derived as the residuals of unadjusted mtDNA-CN regressed against age
(restricted cubic splines with 4 degrees of freedom), sex, log-transformed white blood
cell, red blood cell, lymphocyte, and neutrophil counts (all using restricted cubic splines
with 4 degrees of freedom), platelet counts, and log-transformed eosinophil, basophil,
and nucleated RBC counts. See Supplementary material for details. The residuals are
standardized with mean of 0 and standard deviation of 1.

### mtDNA-CN in Asthmatics in SARP

SARP was designed to characterize severe asthma at the molecular, cellular, and clinical levels and includes detailed clinical and mechanistic data. Thus, this cohort allows investigation of mtDNA-CN in relation to asthma characteristics and mechanisms. The clinical characteristics of the participants of SARP are displayed in Table 3. Compared to UKB, the participants in SARP are younger, have higher BMI, worse lung functions, and higher eosinophil counts. The SARP cohort includes all racial and ethnic backgrounds. In the UKB, severity was defined based on asthma medication use in self-reported asthma; in contrast, in SARP, asthma was validated based upon American Thoracic Society (ATS) guidelines, which include a positive methacholine challenge test and/or reversible airflow obstruction, and asthma severity classified based on ATS and European Respiratory Society (ERS) guidelines (14, 15). As compared to non-severe asthmatics, severe asthmatics in SARP have lower lung functions, higher white blood cell (WBC), neutrophil, and eosinophil counts, and worse asthma control as measured with the Asthma Control Test (Table 3).

**Table 3.** Features of participants in the Severe Asthma Research Program (SARP)

| Characteristics | Non-severe<br>Asthma<br>(*n* = 703) | Severe<br>Asthma<br>(*n* = 580) | *P*<br>Value* |
| --- | --- | --- | --- |
| Age — yr | 37 ± 14 | 47 ± 13 | <0.0001 |
| Female — % | 67 | 66 | 0.6 |
| Ancestry — % |  |  |  |
| European ancestry | 65 | 66 | 0.7 |
| African ancestry | 26 | 26 | 0.9 |
| Body mass index — kg/m<sup>2</sup> | 29.8 ± 7.9 | 32.5 ± 8.7 | <0.0001 |
| Blood pressure — mm Hg |  |  |  |
| Systolic | 122 ± 14 | 128 ± 15 | <.0001 |
| Diastolic | 75 ± 10 | 78 ± 10 | <0.0001 |
| Hemoglobin — g/dL | 14.1 ± 2.5 | 14.7 ± 8.1 | 0.2 |
| Hematocrit — % | 41.1 ± 4.1 | 41.8 ± 5.5 | 0.08 |
| Lung functions |  |  |  |
| Forced expiratory volume in 1 second — %<br>predicted | 82 ± 17 | 64 ± 21 | <0.0001 |
| Forced vital capacity — % predicted | 92 ± 16 | 78 ± 18 | <0.0001 |
| FEV<sub>1</sub>/FVC | 0.74 ± 0.10 | 0.65 ± 0.12 | <0.0001 |
| Asthma control test<sup>†</sup> | 19.4 ± 4.0 | 15.3 ± 4.8 | <0.0001 |
| White blood cells — x10<sup>9</sup>/liter | 6.8 ± 2.1 | 8.0 ± 2.9 | <0.0001 |
| Neutrophils | 3.9 ± 1.7 | 5.0 ± 2.6 | <0.0001 |
| Eosinophils | 0.25 ± 0.21 | 0.29 ± 0.27 | 0.01 |
| IgE — IU/ml | 337 ± 799 | 331 ± 547 | 0.8 |
| Glutathione peroxidase — U/ml | 0.085 ± 0.021 | 0.091 ± 0.025 | 0.05 |
| Superoxide dismutase — U/ml | 18.6 ± 10.9 | 19.7 ± 10.4 | 0.13 |
| Fractional exhaled nitric oxide — ppb | 37 ± 34 | 35 ± 36 | 0.5 |
| Exacerbations in prior year | 0.61 ± 1.19 | 2.37 ± 2.82 | <0.0001 |
| Exacerbations in one-year follow-up | 0.36 ± 0.88 | 1.38 ± 1.98 | <0.0001 |
| mtDNA-CN | 92.0 ± 70.5 | 80.0 ± 64.2 | 0.001 |
| Unadjusted mtDNA-CN residuals | 0.080 ± 1.037 | -0.097 ± 0.945 | 0.001 |
| Adjusted mtDNA-CN | 0.004 ± 1.038 | -0.005 ± 0.953 | 0.8 |

Mean ± SD; \**P* value for non-severe asthmatics vs. severe asthmatics. <sup>†</sup>Scores of 19 or
higher are well controlled.

Variation in mtDNA-CN is observed by sex and ethnicity (Figure E1). After adjusting for age, sex, ethnicity, WBC and other covariates, severe and non-severe asthmatics have similar mtDNA-CN (Table 3). BMI is related to mtDNA-CN among all asthma (*r =* 0.060, *P =* 0.03, *n* = 1237). Linear regression analyses suggests that mtDNA-CN in severe asthma may tend to decline more with age than in non-severe asthma (*P* = 0.09 severe asthmatics *vs*. non-severe asthmatics.) (Figure 3A). However, when mtDNA-CN is adjusted for covariates, age-related decline of mtDNA-CN is no longer observed in this cohort that is smaller in number and relatively younger than the UKB cohort (Figure 3B).

**Figure 3.**
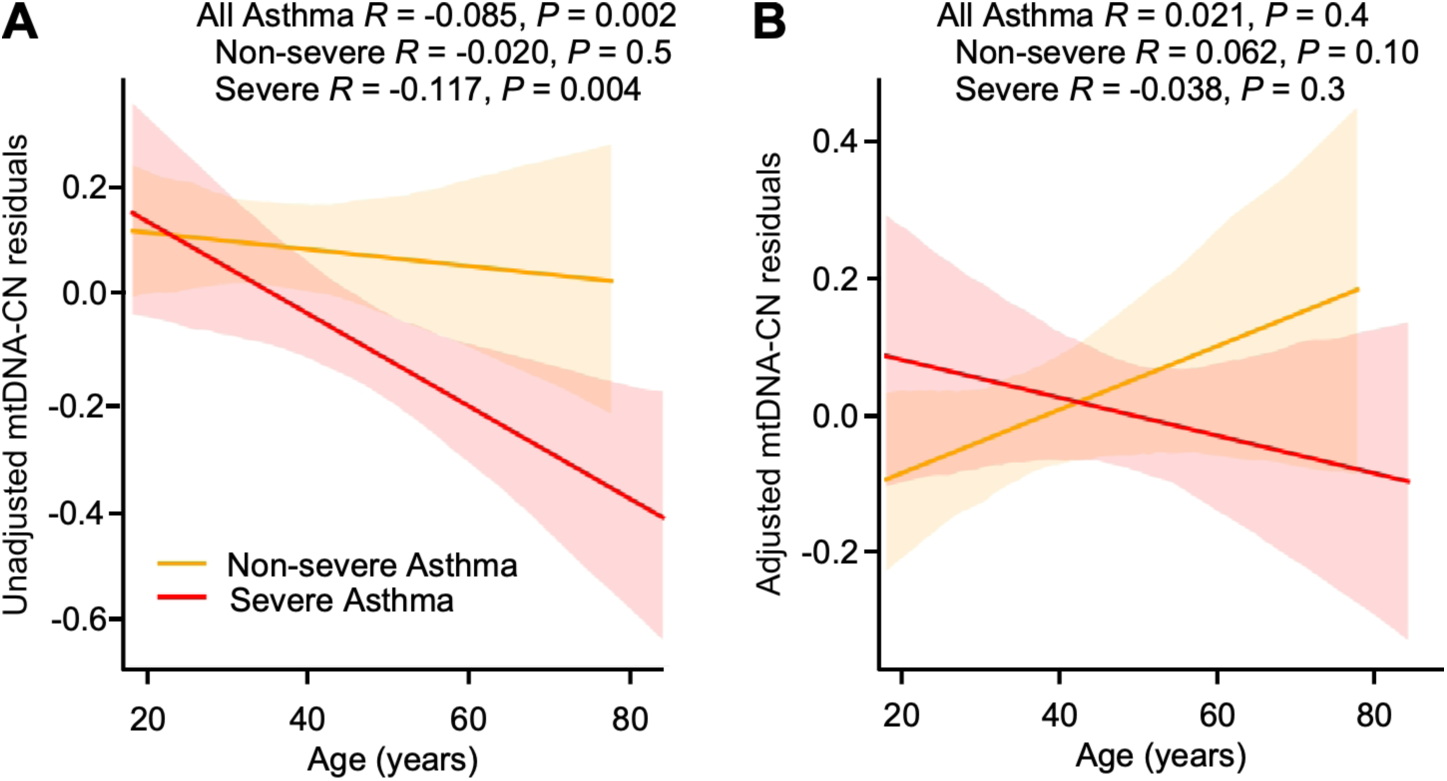
mtDNA-CN related with age in SARP. (*A*) mtDNA-CN residuals are inversely related to age in severe asthmatics and all asthmatics but not in non-severe asthmatics, (*B*) while adjusted mtDNA-CN is not related to age in severe or non-severe asthmatics.

### Increased Oxidative Stress in Asthma of SARP

A small number of healthy individuals without asthma were included in SARP as previously described (Table E3) (20), but these do not have mtDNA-CN available. Here, asthmatics have greater nitrative stress than healthy controls as measured by F_E_NO as previously reported (Table E3) (20, 21). In an early report from SARP with 114 asthmatics and 20 healthy controls (22), asthmatics were reported to have lower serum SOD as compared with controls (4, 5, 18, 22). Similarly here, in the larger SARP cohort, SOD activity is lower in asthma as compared to healthy controls (SOD U/ml, controls 22.3 ± 10.8, *n* = 114, asthmatics 19.1 ± 10.7, *n* = 960; *P* = 0.003) (Table E3), but severe and non-severe asthma have similar SOD (Table 3). GPx is similar among asthmatics and controls, as previously published in a much smaller cohort (22) (GPx U/ml, controls 0.087 ± 0.020, *n* = 44, asthmatics 0.087 ± 0.023, *n* = 297; *P* = 0.9) (Table E3). These results support the paradigm that asthma inflammation is characterized by changes in redox balance as compared to healthy controls (4, 5, 23), but the severe and non-severe groups are similar in the measures of SOD, GPx, F_E_NO, and mtDNA-CN (Table 3).

### mtDNA-CN Predicts Asthma Exacerbations

Predicting and preventing exacerbation frequency is critical for the treatment of asthma (16, 24). Here, participants with asthma in one-year longitudinal follow-up in SARP III (*n* = 468) are analyzed for associations of mtDNA-CN and SOD with exacerbations. Based on previous studies in SARP (16, 25), asthmatics are classified into three groups [0, 1 – 2, or ≥3] by the number of exacerbations in one-year longitudinal follow-up (Table 4). As previously reported (16), of 468 asthmatics, 281 (60%) were without exacerbation in one-year longitudinal follow-up, while 126 (27%) had one or two exacerbations, and 61 (13%) had three or more exacerbations (Table 4). Fifty-five of the 61 (90.2%) asthmatics of the group with three or more exacerbations have severe disease; 131 of the 281 (46.6%) asthmatics of the group with no exacerbations, and 91 of the 126 (72.2%) asthmatics of the group with one or two exacerbations have severe disease (Table 4). As previously reported (16), F_E_NO levels are different among the three groups (ANOVA *P* = 0.01) (Table 4). Here, mtDNA-CN is lowest in the group with three or more exacerbations in longitudinal follow-up (Table 4). In contrast to association with future exacerbations, the mtDNA-CN is not associated with numbers of exacerbations in the prior year [1-2 *vs.* 0 exacerbation group, *P* = 0.2; 3 or more *vs.* 0 exacerbation group, *P* = 0.7), suggesting that high mtDNA-CN levels may be protective from future exacerbations.

**Table 4.**
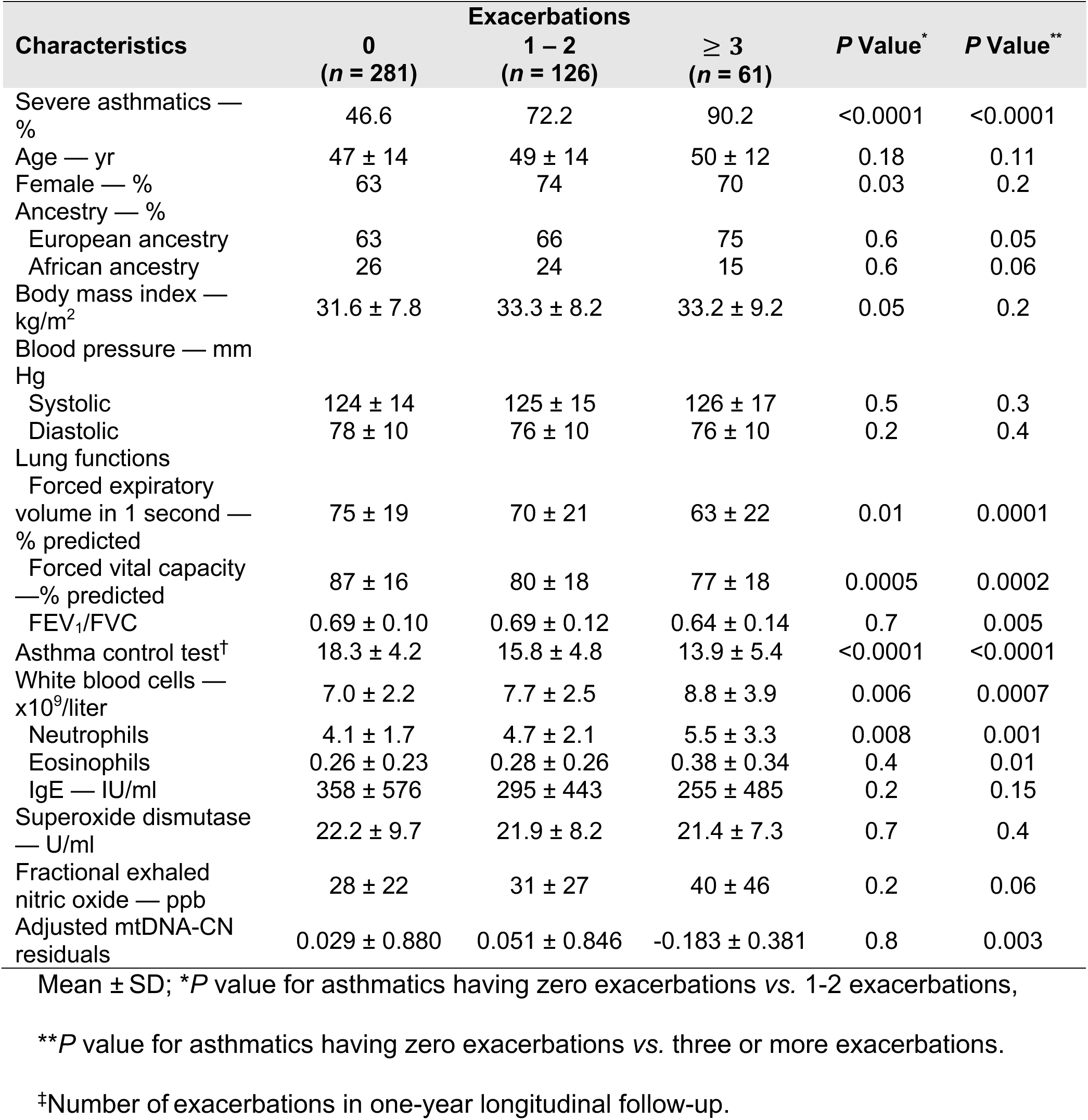
Features of study participants categorized by frequency of exacerbations^‡^ in SARP.

| Characteristics | Exacerbations |  |  | *P* Value* | *P* Value** |
| --- | --- | --- | --- | --- | --- |
|  | 0<br>(*n* = 281) | 1 – 2<br>(*n* = 126) | ≥ 3<br>(*n* = 61) |  |  |
| Severe asthmatics — % | 46.6 | 72.2 | 90.2 | <0.0001 | <0.0001 |
| Age — yr | 47 ± 14 | 49 ± 14 | 50 ± 12 | 0.18 | 0.11 |
| Female — % | 63 | 74 | 70 | 0.03 | 0.2 |
| Ancestry — % |  |  |  |  |  |
| European ancestry | 63 | 66 | 75 | 0.6 | 0.05 |
| African ancestry | 26 | 24 | 15 | 0.6 | 0.06 |
| Body mass index — kg/m<sup>2</sup> | 31.6 ± 7.8 | 33.3 ± 8.2 | 33.2 ± 9.2 | 0.05 | 0.2 |
| Blood pressure — mm Hg |  |  |  |  |  |
| Systolic | 124 ± 14 | 125 ± 15 | 126 ± 17 | 0.5 | 0.3 |
| Diastolic | 78 ± 10 | 76 ± 10 | 76 ± 10 | 0.2 | 0.4 |
| Lung functions |  |  |  |  |  |
| Forced expiratory volume in 1 second — % predicted | 75 ± 19 | 70 ± 21 | 63 ± 22 | 0.01 | 0.0001 |
| Forced vital capacity — % predicted | 87 ± 16 | 80 ± 18 | 77 ± 18 | 0.0005 | 0.0002 |
| FEV<sub>1</sub>/FVC | 0.69 ± 0.10 | 0.69 ± 0.12 | 0.64 ± 0.14 | 0.7 | 0.005 |
| Asthma control test<sup>‡</sup> | 18.3 ± 4.2 | 15.8 ± 4.8 | 13.9 ± 5.4 | <0.0001 | <0.0001 |
| White blood cells — x10<sup>9</sup>/liter | 7.0 ± 2.2 | 7.7 ± 2.5 | 8.8 ± 3.9 | 0.006 | 0.0007 |
| Neutrophils | 4.1 ± 1.7 | 4.7 ± 2.1 | 5.5 ± 3.3 | 0.008 | 0.001 |
| Eosinophils | 0.26 ± 0.23 | 0.28 ± 0.26 | 0.38 ± 0.34 | 0.4 | 0.01 |
| IgE — IU/ml | 358 ± 576 | 295 ± 443 | 255 ± 485 | 0.2 | 0.15 |
| Superoxide dismutase — U/ml | 22.2 ± 9.7 | 21.9 ± 8.2 | 21.4 ± 7.3 | 0.7 | 0.4 |
| Fractional exhaled nitric oxide — ppb | 28 ± 22 | 31 ± 27 | 40 ± 46 | 0.2 | 0.06 |
| Adjusted mtDNA-CN residuals | 0.029 ± 0.880 | 0.051 ± 0.846 | -0.183 ± 0.381 | 0.8 | 0.003 |

Mean ± SD; \**P* value for asthmatics having zero exacerbations vs. 1-2 exacerbations,
\*\**P* value for asthmatics having zero exacerbations vs. three or more exacerbations.
<sup>‡</sup>Number of exacerbations in one-year longitudinal follow-up.

To study the association of mtDNA-CN to the likelihood that asthma exacerbation will occur, odds ratios (OR) are calculated. In evaluation of mtDNA quartiles, the top quartile of mtDNA-CN has significantly lower odds (OR 0.257 [95% CI, 0.099 to 1.670], *P* = 0.005) of three or more exacerbations as compared with the bottom quartile (Figure E2). The top-quartile mtDNA-CN individuals have features similar to the bottom-quartile, including the numbers that have severe asthma, but differ in having a higher BMI (*P* = 0.005) (Table E4).

To determine cutoff values of mtDNA, the dichotomized biomarker at the cutoffs e.g., -0.033 cutoff for mtDNA, are examined to analyze the odds of having three or more exacerbations *vs*. zero exacerbations (Table E5). Asthmatics with mtDNA-CN below the cutoff are older than those with mtDNA-CN above the cutoff (*P* = 0.04) (Table E6). The odd ratio of mtDNA-CN is 0.352 ([95% CI, 0.164 to 0.753], *P* = 0.007) (Figure 4), indicating that asthmatics with levels of mtDNA-CN above the cutoff have ∼35% of the risk of three or more exacerbations compared to those with mtDNA-CN levels below the cutoff. Similarly, using continuous analyses per SD increase, mtDNA-CN predicts those who will have few exacerbations (OR 0.442 [95% CI, 0.224 to 0.875], *P* = 0.01), while F_E_NO predicts those who will have more exacerbations (OR 1.387 [95% CI, 1.090 to 1.764], *P* = 0.008) (Figure E3B). F_E_NO levels (higher than cutoff of 27.5 ppb) increase exacerbation risk twofold (*P =* 0.009) (Figure 4) as previously reported (16, 21). SOD is not predictive of exacerbations.

**Figure 4.**
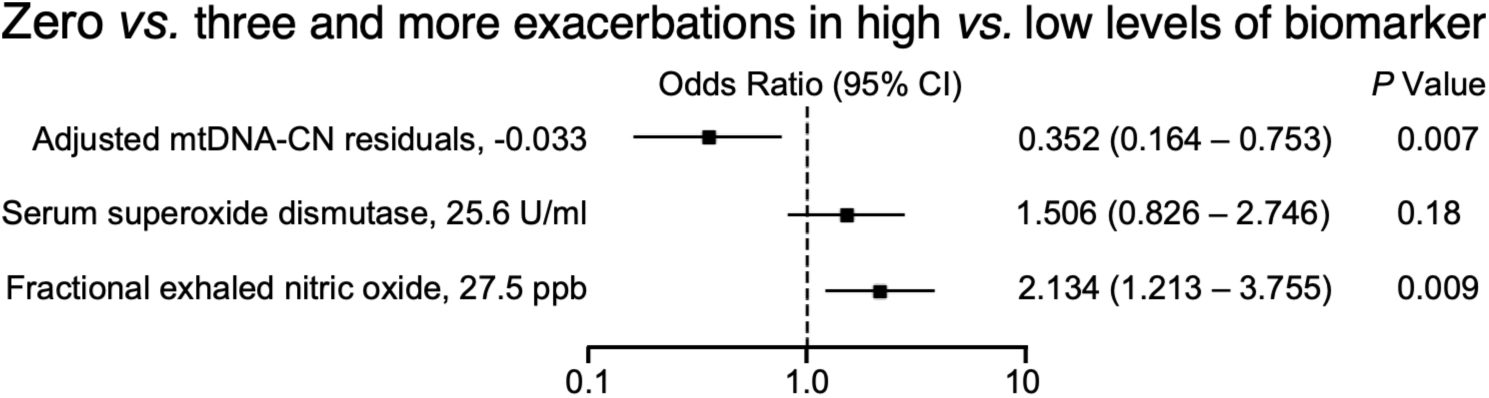
Odds ratios for risk of three or more exacerbations in the subsequent year for biomarkers of asthma. Cutoff values were determined (Table E5) for mtDNA-CN, serum superoxide dismutase activity, and fractional exhaled nitric oxide. A higher mtDNA-CN predicted a lower risk of three or more exacerbations, while a higher fractional exhaled nitric oxide predicted a greater risk of three or more exacerbations.

## Discussion

This study shows that asthma is associated with low mtDNA-CN, an indicator of mitochondrial dysfunction. Interestingly, although asthmatics have lower mtDNA-CN as compared to non-asthmatics, the asthma severity groups did not differ with respect to mtDNA-CN. However, higher mtDNA-CN predicts less frequent exacerbations in one-year follow-up, suggesting mitochondrial dysfunction contributes to the exacerbation-prone asthma phenotype.

Mitochondria are principal producers of ROS and regulate redox status in the cell. Due to multiple copies of mtDNA, lack of a protective histone protein, and limited mtDNA repair capacity, mtDNA is also more susceptible to damage by excess ROS, which may lead to dysregulation of the respiratory chain and further increase ROS production (26-30). The non-coding regulatory D-loop region (mtDNA control region) is known to be more sensitive to ROS-mediated damage than the coding portion of the mitochondrial genome and the nuclear genome (31, 32). Increased mtDNA D-loop methylation induced by excess ROS was associated with reduced mtDNA-CN (33). In the Study of Asthma Phenotypes and Pharmacogenomic Interactions by Race-Ethnicity (SAPPHIRE), mtDNA-CN in whole blood was higher in African American asthmatics as compared with their non-asthmatic counterparts, but mtDNA-CN was not related to asthma severity (34). Here, we also found mtDNA-CN is not significantly different between severe and non-severe asthma after adjusting for age, sex, ethnicity, and blood cell counts in both UKB and SARP cohorts; however, asthmatics, regardless of severity, have significantly lower mtDNA-CN as compared with non-asthmatics. The UKB and SARP cohorts in this study differ from the SAPPHIRE cohort in genetic ancestries, ages, smoking status, BMI, WBC counts, and sample size of studies, all of which might lead to the discrepant findings. Here, asthmatics with African ancestry have higher levels than those with European ancestry. Further studies are needed to understand the influence of ancestry and mitochondrial function.

Previous studies indicated that mtDNA-CN is affected by and should be adjusted for age (35-38). Our results show that mtDNA-CN is inversely related to age in all asthmatics but does not account for differences among asthmatics and non-asthmatics. Asthmatics have lower mtDNA-CN at every age in comparison to non-asthmatics. The mtDNA-CN per cell is estimated from genomic DNA derived from buffy coat, which greatly depends on peripheral blood cell type, platelet count, and WBC counts (35, 39, 40). Previously, others reported that mtDNA-CN was inversely correlated with WBC count in their study populations (35, 39, 40). Here, WBC and neutrophil counts are significantly higher in severe asthmatics than in non-severe asthmatics and inversely related with mtDNA-CN. In SARP, blood eosinophil counts are higher in severe asthmatics than in non-severe asthmatics but are not related with mtDNA-CN, indicating that lower levels of mtDNA-CN may be related to neutrophilic inflammation and not Th2 inflammation. Mitochondria numbers can vary in different cells and/or tissues. Prior work has shown changes in mitochondrial function in platelets and airway epithelial cells from asthmatics (41). Interestingly, although asthma is associated with obesity (42), resting energy expenditure is elevated in asthmatics as compared to non-asthmatics (43), supporting the concept of changes in mitochondrial function.

Asthma is a common problem. Severe asthma consumes a great deal of resources. However, there are very few biomarkers at the molecular level that discriminate phenotypes of asthma, relative risks for asthma severity, and/or risks of frequent exacerbations. Previous findings showed that F_E_NO (44, 45), blood eosinophils (16, 24, 45, 46), sputum eosinophils (16, 44), body mass index (16, 47), age (16, 47), and urinary total conjugated 3-bromotyrosine (48) are associated with uncontrolled asthma and exacerbation frequency. Existing biomarkers can be grouped into categories, e.g., related to redox pathways, arginine/NO pathway, and/or Th2 eosinophilic pathway (16, 24, 44-47, 49, 50). Although frequent exacerbations happen in a small number of patients, prediction of asthma exacerbations is important for optimal care of individuals (16, 24). Prior reports in SARP cohort indicate that blood eosinophils, BMI, and bronchodilator responsiveness were associated with exacerbation frequency in the prior year (16); high plasma IL-6 and blood eosinophils were associated with increased incident rate of exacerbations over three years of follow-up (24), and urinary total conjugated 3-bromotyrosine predicted exacerbation risk over one year of follow-up (48). Here, asthmatics with high mtDNA-CN levels have a 65% reduced risk of having three or more exacerbations than those with lower mtDNA-CN. Unlike other measures, we did not find any association of mtDNA-CN to prior year exacerbations. This may suggest that changes in mtDNA-CN occur in advance of exacerbations. The loss of SOD activity in asthma is well described and related to oxidative inactivation due to cleavage of the C57-C146 disulfide bond and exposure of usually unavailable cysteines (6). However, loss of SOD activity is not associated with exacerbations. A prior study suggests a relationship of SOD to severity of airflow obstruction (22).

Overall, this study identifies that mitochondrial dysfunction is present in asthma and may play a role in the molecular basis of asthma and exacerbations. Higher mtDNA-CN, an indicator of better mitochondrial function, is predictive of fewer future exacerbations, suggesting that targeting mitochondria and/or metabolism may be useful in future approaches for treatment of exacerbation-prone asthma.

## Data Availability

All data produced in the present study are available upon reasonable request to the authors.

## Acknowledgments

We gratefully acknowledge the studies and participants who provided biological samples and data for SARP, UKB, and TOPMed. The authors would like to thank SARP clinical coordinators and laboratory personnel for their contribution to fulfilling SARP’s scientific mission. The research findings resulting from SARP would not have been possible without their dedication and assistance with study visits, data collection, and biological sample processing. The authors also thank Peter Bazeley, Thomas W. Blackwell, Xiaoqi Geng, Goncalo Abecasis, Jun Ding for mitochondrial DNA copy number estimation.

SARP3 acknowledgement (industry): The following companies provided financial support for study activities at the Coordinating and Clinical Centers beyond the third year of patient follow-up: AstraZeneca, Boehringer-Ingelheim, Genentech, GlaxoSmithKline, Sanofi–Genzyme–Regeneron, and TEVA.

