## Supplement for "Mitochondrial DNA Copy Number Variation in Asthma Risk, Severity, and Exacerbations"

Table of Contents

1. UKB dataset, methods, and supplementary Tables E1 and E2……………….…………3
2. SARP dataset……………………………………………………………………….9
3. Adjusted mtDNA-CN and analysis…….……………………………………..…10
4. Supplementary Tables E3 – E6………………………........................……….11
5. Supplementary Figures E1 – E2……………..……………………………………...15

**Supplementary Methods**

**Supplementary UKB Dataset Methods**

**I. UKB curated dataset criteria**

The UK Biobank (UKB) dataset includes 502,485 participants (54.4% female, 12.1% prevalent asthma) with age range 37–73 years (mean ± SD, Male 56 ± 8, Female 56 ± 8) recruited between 2006 and 2010 throughout the UK across 22 assessment centers. Exclusion reasons were non-Whites (N = 28,910), related individuals (N = 78,660), cell count missing (N = 12,022), cell count outliers (N = 3,667), exclusion criteria (N = 96,787), without medication information (*n* = 53), and those who were not genotyped (N = 13,460)

Inclusion criteria. We include asthmatics and controls (non-asthmatic).

Cell count outliers are excluded due to known influence on mtDNA-CN (E1). For mtDNA-CN analyses, we excluded those who are outliers by cell counts as shown below as previously described (E1).

Cell count and platelets volume and size outliers were excluded.

| Excluded Outliers |
|---|
| Log(WBC) ≤ 1.25 or ≥ 3 [WBC ≤ 3.49 or ≥ 20.08 K/ul] |
| Log(RBC) ≤ 1.4 or ≥ 2 [RBC ≤ 4.0 or ≥ 7.3 x$10^6$/ul] |
| Platelet ≤ 10 or ≥ 500 |
| Log(Lymphocyte) ≤ 0.10 or ≥ 2 [Lymphocyte ≤ 1.1 or ≥ 7.3 K/ul] |
| Log(Mono) ≥ 0.9 [Mono ≥ 2.4 K/ul] |
| Log(Neutrophil) ≤ 0.75 or ≥ 2.75 [Neutrophil ≤ 2.1 or ≥ 15.6 K/ul] |
| Log(Eos) ≥ 0.75 [Eos ≥ 2.1 K/ul] |
| Log(Baso) ≥ 0.45 [Baso ≥ 1.5 K/ul] |

Exclusion criteria specific to this study:

a. Exclusion criteria:
    1. Current smokers
    2. Prevalent cancer
    3. Prevalent chronic obstructive pulmonary disease (COPD)
    4. Cystic fibrosis (CF)
    5. Gastro-oesophageal reflux disease (GORD, GERD)
    6. Chronic sinusitis
    7. Sleep apnea
    8. Renal/kidney failure
    9. Hepatic failure
    10. Heart failure

**b.** In the UKB to find these to exclude, we used ICD9 code(s), ICD10 code(s), self-report code(s), and Data-Field(s) (see **Table E1**).

**Table E1.** UKB ICD9 code(s), the ICD10 code(s), self-report code(s) and Data-Field(s) of exclusion criteria*.

| # | Comorbidity | ICD9 | ICD10 | Self-Reported Codes | Data-Field |
|---|---|---|---|---|---|
| **1** | **Current smokers** | Not reported | | Not reported | 20116 Smoking status (*n*=55687) |
| **2** | **Prevalent cancer** | Not reported | Not reported | Not reported | 40005 Date of cancer diagnosis before study entry date (field: 53) (*n*=89765) |
| **3** | **COPD** | | | | |
| **4** | **CF** | Not reported | E84.0 Cystic fibrosis with pulmonary manifestations (*n*=16)<br>J84.0 Alveolar and parietoalveolar conditions (*n*=22)<br>J84.1 Other interstitial pulmonary diseases with fibrosis (*n*=2703)<br>J84.8 Other specified interstitial pulmonary diseases (*n*=115) | 1121 pulmonary fibrosis<br>1115 interstitial lung disease<br>1122 fibrosing alveolitis/unspecified alveolitis | |

| | | | | | |
|---|---|---|---|---|---|
| | | | J84.9 Interstitial pulmonary disease, unspecified (*n*=958) | | |
| 5 | **GERD** | 53010 Oesophageal reflux without mention of oesophagitis (*n*=8)<br>53011 Oesophagitis (with oesophageal reflux) (*n*=12) | K21.0 Gastro-oesophageal reflux disease with oesophagitis (*n*=18782)<br>K21.9 Gastro-oesophageal reflux disease without oesophagitis (*n*=37933) | 1138 gastro-oesophageal reflux (gord) / gastric reflux | |
| 6 | **Chronic Sinusitis** | 4730 Chronic sinusitis, maxillary (*n*=34)<br>4731 Chronic sinusitis, frontal (*n*=5)<br>4732 Chronic sinusitis, ethmoidal (*n*=13)<br>4733 Chronic sinusitis, sphenoidal (*n*=1)<br>4738 Other specified chronic sinusitis (*n*=2)<br>4739 Chronic sinusitis, unspecified (*n*=185) | J32.0 Chronic maxillary sinusitis (*n*=778)<br>J32.1 Chronic frontal sinusitis (*n*=73)<br>J32.2 Chronic ethmoidal sinusitis (*n*=216)<br>J32.3 Chronic sphenoidal sinusitis (*n*=45)<br>J32.4 Chronic pansinusitis (*n*=186)<br>J32.8 Other chronic sinusitis (*n*=263)<br>J32.9 Chronic sinusitis, unspecified (*n*=3017) | 1416 chronic sinusitis | |
| 7 | **Sleep apnea** | Not reported | G47.3 Sleep apnea (*n*=9647) | 1123 sleep apnea | |
| 8 | **Renal/kidney failure** | 5859 Chronic renal failure (*n*=18)<br>5869 Renal failure, unspecified (*n*=9) | N18.0 End-stage renal disease (*n*=461)<br>N18.1 Chronic kidney disease, stage 1 (*n*=187)<br>N18.2 Chronic kidney disease, stage 2 (*n*=1350)<br>N18.3 Chronic kidney disease, stage 3 (*n*=10462)<br>N18.4 Chronic kidney disease, stage 4 (*n*=1487)<br>N18.5 Chronic kidney disease, stage 5 (*n*=1248)<br>N18.8 Other chronic renal failure (*n*=73)<br>N18.9 Chronic renal failure, unspecified (*n*=9095)<br>N19 Unspecified renal failure (*n*=2542) | 1192 renal/kidney failure<br>1193 renal failure requiring dialysis<br>1194 renal failure not requiring dialysis | ESRD as defined by UKB. (*n*=787) |
| 9 | **Hepatic failure** | 5712 Alcoholic cirrhosis of liver (*n*=1)<br>5714 Chronic hepatitis (*n*=13)<br>5715 Cirrhosis of liver without mention of alcohol (*n*=4)<br>5716 Biliary cirrhosis (*n*=7) | K72.0 Acute and subacute hepatic failure (*n*=246)<br>K72.1 Chronic hepatic failure (*n*=39)<br>K72.9 Hepatic failure, unspecified (*n*=739)<br>K73.0 Chronic persistent hepatitis, not elsewhere classified (*n*=2)<br>K73.1 Chronic lobular hepatitis, not elsewhere classified (*n*=9) | 1158 liver failure/cirrhosis | |

| | | | | |
|---|---|---|---|---|
| | | | K73.2 Chronic active hepatitis, not elsewhere classified (*n*=76)<br>K73.8 Other chronic hepatitis, not elsewhere classified (*n*=22)<br>K73.9 Chronic hepatitis, unspecified (*n*=149)<br>K74.0 Hepatic fibrosis (*n*=331)<br>K74.1 Hepatic sclerosis (*n*=29)<br>K74.2 Hepatic fibrosis with hepatic sclerosis (*n*=7)<br>K74.3 Primary biliary cirrhosis (*n*=393)<br>K74.4 Secondary biliary cirrhosis (*n*=24)<br>K74.5 Biliary cirrhosis, unspecified (*n*=134)<br>K74.6 Other and unspecified cirrhosis of liver (*n*=1550) | |
| **10** | **Heart failure** | 4280 Congestive heart failure (*n*=6)<br>4281 Left heart failure (*n*=13)<br>4289 Heart failure, unspecified (*n*=2) | I50.0 Congestive heart failure (*n*=6650)<br>I50.1 Left ventricular failure (*n*=8432)<br>I50.9 Heart failure, unspecified (*n*=5701) | 1076 heart failure/pulmonary odema |

*Count (*n*) for each item is included.

### II. <u>UKB lung function methods</u>

Because many participants do not have calculated FEV1 % predicted, we are inputting predicted lung functions so we can analyze % FEV1. See **Table E2** below for coefficients from prediction equations and R Codes to calculate FEV1 and FVC percent predicted (E2) provided by Joe Zein**.**

**Table E2.** Coefficients from prediction equations for normal lung function measures by sex.

| | $b_0$ | $b_1$ | $b_2$ | $b_3$ | |
|---|---|---|---|---|---|
| Corresponding variable | Intercept | A | $A^2$ | ln H | **Equation** |
| **ln $FEV_1$ L** | | | | | |
| Males ($R^2$=0.59; σ=0.14) | | | | | |
| Mean | | | | | |
| Age ≤25 yrs | -10.41186 | 0.09569 | -0.00221 | 2.10839 | $e^{-10.41186 + 0.09569A -0.00221A^2 + 2.10839 \ln H}$ |
| Age >25 yrs | -9.37674 | 0.00183 | -0.00011 | 2.10839 | $e^{-9.37674 + 0.00183A - 0.00011A^2 + 2.10839 \ln H}$ |
| LLN | | | | | |
| Age ≤25 yrs | -10.75820 | 0.10320 | -0.00231 | 2.10839 | $e^{-10.75820 + 0.10320A - 0.00231A^2 + 2.10839 \ln H}$ |
| Age >25 yrs | -9.72308 | 0.00933 | -0.00021 | 2.10839 | $e^{-9.72308 + 0.00933A - 0.00021A^2 + 2.10839 \ln H}$ |
| Females ($R^2$=0.69; σ=0.14) | | | | | |
| Mean | -8.49717 | 0.00422 | -0.00015 | 1.90019 | $e^{-8.49717 + 0.00422A - 0.00015A^2 + 1.90019 \ln H}$ |
| LLN | -8.68467 | 0.00495 | -0.00018 | 1.90019 | $e^{-8.68467 + 0.00495A - 0.00018A^2 + 1.90019 \ln H}$ |
| **ln FVC L** | | | | | |
| Males ($R^2$=0.52; σ=0.14) | | | | | |
| Mean | | | | | |
| Age ≤25 yrs | -11.45146 | 0.09895 | -0.00216 | 2.32222 | $e^{-11.45146 + 0.09895A - 0.00216A^2 + 2.32222 \ln H}$ |
| Age >25 yrs | -10.36706 | 0.00434 | -0.00011 | 2.32222 | $e^{-10.36706 + 0.00434A - 0.00011A^2 +2.32222 \ln H}$ |
| LLN | | | | | |
| Age ≤25 yrs | -11.63230 | 0.09795 | -0.00217 | 2.32222 | $e^{-11.63230 + 0.09795A - 0.00217A^2 + 2.32222 \ln H}$ |
| Age >25 yrs | -10.54790 | 0.00334 | -0.00012 | 2.32222 | $e^{-10.54790 + 0.00334A - 0.00012A^2 + 2.32222 \ln H}$ |
| Females ($R^2$=0.62; σ=0.15) | | | | | |
| Mean | -9.66999 | 0.00837 | -0.00017 | 2.14118 | $e^{-9.66999 + 0.00837A - 0.00017A^2 + 2.14118 \ln H}$ |
| LLN | -9.84941 | 0.00772 | -0.00018 | 2.14118 | $e^{-9.84941 + 0.00772A -0.00018A^2 + 2.14118 \ln H}$ |
| **ln FEV1/FVC** | | | | | |

| | | | | | |
|---|---|---|---|---|---|
| Males ($R^2$=0.15; σ=0.08) | | | | | |
| Mean | 1.03981 | -0.00394 | 0.00002 | -0.21653 | $e^{1.03981 - 0.00394A + 0.00002A\text{^}2 - 0.21653\ln H}$ |
| LLN | 0.82621 | 0.00101 | -0.00005 | -0.21653 | $e^{0.82621+ 0.00101A - 0.00005A\text{^}2 - 0.21653\ln H}$ |
| Females ($R^2$=0.16; σ=0.08) | | | | | |
| Mean | 1.15822 | -0.00415 | 0.00002 | -0.23815 | $e^{1.15822 - 0.00415A + 0.00002A\text{^}2 - 0.23815\ln H}$ |
| LLN | 1.00699 | -0.00196 | -0.00001 | -0.23815 | $e^{1.00699 - 0.00196A - 0.00001A\text{^}2 -0.23815\ln H}$ |

Model: mean lung function=$e^{b0+b1A+b2A\text{^}2+b3\ln H}$.
$R^2$: fraction of explained variance; σ: SD of residuals; LLN: lower limit of normal; A: age; H: height.

**A. Codes to extract percent predicted for FEV1 in the UKB.**

Please find the codes
Field 3063 and 3062 have 3 measurements that are used to calculate ATS reproducibility

Data-Field 3063
Description: Forced expiratory volume in 1-second (FEV1)

Data-Field 20150 = “FEV1”
Description: Forced expiratory volume in 1-second (FEV1), Best measure

Data-Field 3062
Description: Forced vital capacity (FVC)

Data-Field 20151 = “FVC”
Description: Forced vital capacity (FVC), Best measure

Data-Field 20152
Description: Reproducibility of spirometry measurement using ERS/ATS criteria

Data-Field 21003
Description: Age when attended assessment center

Data-Field 21022
Description: Age at recruitment

Data-Field 50
Description: Standing height

Data-Field 31
Description: Sex

Data-Field 21000
Description: Ethnic background

Data-Field 20255
Description: Spirometry QC measure

B. **R Codes to calculate FEV1 and FVC percent predicted**.

```
#1– Step 1: Define FEV1
######################

#Merging all fev1 measurements
FEV1_merged <- cbind(uk.b$Forced_expiratory_volume_in_1_second_FEV1,
uk.b$Forced_expiratory_volume_in_1_second_FEV1_2,
                uk.b$Forced_expiratory_volume_in_1_second_FEV1_3)
dim(FEV1_merged)

# function to identify max FEV1
uk.b$FEV1_max <- apply(FEV1_merged, 1, function(x){
  max(x,na.rm = T)
})

#change -Inf to NA
uk.b$FEV1_max[uk.b$FEV1_max == -Inf] <- NA


#2 – Step 2: Define FVC max
######################
```

```
# merging all fvc measurements
FVC_merged <- cbind(uk.b$Forced_vital_capacity_FVC, uk.b$Forced_vital_capacity_FVC_2,
             uk.b$Forced_vital_capacity_FVC_3)
dim(FVC_merged)

# function to identify max FVC
uk.b$FVC_max <- apply(FVC_merged, 1, function(x){
  max(x,na.rm = T)
})

#change -Inf to NA
uk.b$FVC_max[uk.b$FVC_max == -Inf] <- NA
head(FVC_merged,10)

#dput(names(uk))


#3 – Step 3: Define Predicted FEV1
########################

#   FEV1 Predicted values calculated from max 3 visits according to UKB formula
uk$FEV1pred<- exp(ifelse(uk$Sex ==1 & uk$age <=25, (-10.41186 + 0.09569*uk$age - 0.00221*uk$age*uk$age +
(2.10839*(log(uk$Standing_height)))),
                  ifelse(uk$Sex ==1 & uk$age > 25, (-9.37674 + 0.00183*uk$age - 0.00011*uk$age*uk$age +
(2.10839*(log(uk$Standing_height)))),
                         ifelse(uk$Sex ==0, (-8.49717 + 0.00422*uk$age - 0.00015*uk$age*uk$age +
(1.90019*(log(uk$Standing_height)))),
                                NA))))
describe(uk$FEV1pred)

# Percent predicted from MAXIMUM FEV1 Calculated
uk$fev1pp_cal <- round((uk$FEV1_max/uk$FEV1pred*100), 2)
uk$fev1pp_cal[1:10]


#4 – Step 4: Define Predicted FVC
########################

uk$FVCpred<- exp(ifelse(uk$Sex ==1 & uk$age <=25, (-11.45146 + 0.09895*uk$age - 0.00216*uk$age*uk$age +
(2.32222*(log(uk$Standing_height)))),
                  ifelse(uk$Sex ==1 & uk$age > 25, (-10.36706 + 0.00434*uk$age - 0.00011*uk$age*uk$age +
(2.32222*(log(uk$Standing_height)))),
                         ifelse(uk$Sex ==0, (-9.66999 + 0.00837*uk$age - 0.00017*uk$age*uk$age +
(2.14118*(log(uk$Standing_height)))),
                                NA))))
describe(uk$FVCpred)

## Percent predicted from MAXIMUM FVC Calculated
uk$fvcpp_cal <- round((uk$FVC_max/uk$FVCpred*100), 2)
uk$fvcpp_cal[1:10]
```

### III. <u>Define mild, moderate-severe asthma in the UKB</u>.

Asthmatics were defined based on self-report. Moderate-severe asthma or mild asthma was defined by asthma medication use based on the UKB id codes among self-reported asthma patients (E3).

The definition for asthma and severity

a. For a binary yes/no
   i. Asthma: self-reported asthma regardless of use of asthma-related medication
   ii. Non-asthma: absence of self-reported asthma regardless of use of asthma-related medication
b. For asthma severity
   i. Moderate-severe asthma: self-reported asthma AND on medication for moderate-severe asthma (see below)
   ii. Mild asthma: self-reported asthma AND on any medication for asthma, excluding medication for moderate-severe asthma
   iii. Non-asthma: absence of self-reported asthma regardless of use of asthma-related medication (so even if he/she is on medication for mild or severe asthma, this person would be defined as not having asthma)

Medications identifying moderate-severe asthma including the UKB id code (E3).

1140883548 ipratropium
1141182628 tiotropium
1140855320 isoetharine hydrochloride*
1140855322 numotac 10mg m/r tablet
1140855328 aleudrin 20mg tablet**
1140855330 aleudrin 1% spray for nebuliser**
1140855332 iso-autohaler 80micrograms inhaler**
1140855358 theodrox tablet
1140855360 sabidal sr-270 424mg m/r tablet
1140855366 pro-vent 300mg m/r capsule
1140855400 bronchodil 20mg tablet
1140855424 theograd 350mg m/r tablet
1140855426 biophylline 350mg m/r tablet
1140855442 adrenaline+atropine compound spray
1140855496 alupent expectorant 20mg tablet*
1140855504 bricanyl compound tablet
1140855506 bricanyl expectorant elixir
1140855530 nethaprin dospan m/r tablet
1140855540 tedral tablet
1140855542 tedral elixir
1140855540 tedral tablet
1140855542 tedral elixir
1140862092 maxivent 100micrograms inhaler
1140862144 salmeterol
1140862148 serevent 25mcg inhaler
1140862162 bricanyl 5mg tablet
1140862168 bricanyl 250mcg inhaler
1140862222 brelomax 2mg tablet
1140862224 respacal 2mg tablet
1140862236 atrovent 20micrograms inhaler
1140862260 aminophylline
1140862266 phyllocontin continus 225mg m/r tablet
1140862274 pecram 225mg m/r tablet
1140862280 bambec 10mg tablet
1140862290 adrenaline product
1140862292 medihaler-epi 280micrograms inhaler
1140862294 adrenaline acid tartrate 280micrograms inhaler
1140862306 cam 4mg/5ml s/f mixture
1140862310 medihaler-iso 80micrograms inhaler
1140862320 alupent 20mg tablet*
1140862336 amnivent 225 sr m/r tablet
1140862346 choline theophyllinate
1140862348 choledyl 100mg tablet
1140862362 duovent inhaler
1140862364 franol tablet
1140862374 ephedrine hydrochloride+theophylline 11mg/120mg tablet
1140862412 theophylline product
1140862414 nuelin 125mg tablet
1140862418 lasma 300mg m/r tablet
1140862424 slo-phyllin 60mg m/r capsule
1140862432 theo-dur 200mg m/r tablet
1140862438 uniphyllin continus 200mg m/r tablet
1140862474 aerobec 50mcg autohaler
1140868364 prednisone
1140868370 decortisyl 5mg tablet
1140881862 oxivent 100mcg inhaler
1141157264 salmeterol product
1141157402 prednisolone product
1141173346 cortisone
1141176832 seretide 50 evohaler
1141182632 spiriva 18micrograms inhalation capsule
1141195224 formoterol
1141195232 budesonide+formoterol
1141157126 montelukast product
1141157132 singulair 10mg tablet
1141168340 zafirlukast
1141168344 accolate 20mg tablet

**Supplementary SARP Dataset Methods**

**Asthmatic participants**

1882 samples with sequencing data were from Trans-Omics for Precision Medicine (TOPMed). We include both non-severe and severe asthmatics. Exclusion criteria of SARP study of TOPMed included having any of the following: current smokers (more than 5-10 pack years depending on age), other respiratory diseases (e.g., cystic fibrosis or chronic obstructive pulmonary disease), premature birth before 35 weeks' gestation, clinically relevant or untreated gastroesophageal reflux, recurrent sinopulmonary infections or obstructive sleep apnea, and cancer diagnosis in the past five years. Exclusion reasons for this study were without matched IDs in the phenotype dataset (N = 144), without severity information (N = 151), missing one row of DNA position (N = 1), duplicate subjects (N = 2), healthy controls (N = 5), age less than 18 years (N = 291), and blood cell count outlier (N = 5).

For SARP participants, asthma was verified based upon American Thoracic Society guidelines, which include positive methacholine challenge test and/or reversible airflow obstruction. The severity of asthma was classified as severe and non-severe in SARP I & II according to the initial ATS workshop definition of severe asthma (E4) and in SARP III based on American Thoracic Society (ATS) and European Respiratory Society (ERS) guidelines (E5).

**I. <u>SARP dataset cleaning</u>**

A. Inclusion criteria. We include asthmatics including non-severe and severe asthmatics.

B. Exclusion criteria of SARP study of TOPMed:
   a. Current smokers (more than 5-10 pack years depending on age)
   b. Cancer diagnosis in the past five years
   c. Other respiratory diseases [e.g., cystic fibrosis (CF) or chronic obstructive pulmonary disease (COPD)]
   d. Premature birth before 35 weeks' gestation
   e. Clinically relevant or untreated gastroesophageal reflux
   f. Recurrent sinopulmonary infections
   g. Obstructive sleep apnea

C. Exclusion reasons for this study:
   a. Without matched IDs in the phenotype dataset (N = 144)
   b. Without severity information (N = 151)
   c. Missing one row of DNA position (N = 1)
   d. Duplicate subjects (N = 2)
   e. Healthy controls (N = 5)
   f. Age less than 18 years (N = 291)
   g. Five cell count outliers are identified using the IQR method or are excessively high or low based on clinical knowledge ( WBC > 20.08 or < 3.49 $x10^9$/liter; platelet > 500 or < 10 $x10^9$/liter; lymphocyte > 7.3 or < 1.1 $x10^9$/liter; monocyte > 2.4 $x10^9$/liter; neutrophil > 15.6 or < 2.1 $x10^9$/liter; basophil > 1.5 $x10^9$/liter; eosinophil > 2.1 $x10^9$/liter), which including 3 EA and 2 AA (all females, all severe asthmatics, and age range 29–54 years), and from 4 research centers, 2 of SARP I&II and 3 of SARP III

D. Participants include 1283 asthmatics (66.3% female, 703 non-severe asthmatics, and 580 severe asthmatics) with age range 18–84 years (mean ± SD, Male 41 ± 14, Female 41 ± 14), who self-identify as European ancestry (EA, N = 838), African ancestry (AA, N = 339) or other ancestries (American Indian, Asian, Hawaiian and others, N = 106). SARP participants enrolled at 11 clinical research centers including 778 from SARP I & II and 505 from SARP III.

**II. <u>Creating the dataset of SARP.</u>**

FEV1 % predicted is ready to use in SARP.

**Healthy control participants**

Healthy control participants were enrolled in SARP I and II as a comparison group. All healthy control participants were over 18 years old, non-smokers, free of respiratory symptoms, and had normal spirometry, a negative methacholine challenge, and $F_ENO$ < 50 ppb (E6). Any participants with measured $F_ENO$, SOD activity, or GPx activity were included in this analysis (Table E3). Two individuals were excluded (one SOD outlier, one $F_ENO$ outlier).

**Adjusted mtDNA-CN and Analysis**

Because there is variation in mtDNA-CN methodology (Affymetrix Axiom array in the UKB as opposed to mtDNA/nDNA in TOPMed) and because depending on study, mtDNA-CNs and age, sex, and WBC counts measurement can vary, we evaluated the mtDNA-CN residual of each individual relative to the total population mean adjusting for confounding factors.

For the UKB data, as described previously (E1), given the known impact of age, sex, and cell counts on mtDNA-CN, we first used visual inspection to identify outliers for cell counts. We then excluded non-Whites, related individuals (used.in.pca.calculation=0), and cell count outliers and then adjusted for age, sex, and cell counts using a backwards regression, starting with a natural spline (df=4) for each covariate. The final model obtained was ("log_" indicates log-transformed variable):

$$Unadjusted\ \mathrm{mtDNA}_{CN} = \mathrm{ns(age, df = 4)} + \mathrm{sex} + \mathrm{ns(log\_WBC, df = 4)} + \mathrm{ns(log\_RBC, df = 4)} + \mathrm{ns(Platelet, df = 4)} + \mathrm{ns(log\_Lymph, df = 4)} + \mathrm{ns(log\_Neutrophil, df = 4)} + \mathrm{log\_Eos} + \mathrm{log\_Baso} + \mathrm{log\_NucRBC}$$

Beta estimates from these analyses were then used to generate fitted values in the full UKB dataset using the 'predict' function. Adjusted mtDNA CN was derived as the residuals from the above model. For all analyses, the adjusted mtDNA-CN was further standardized by subtracting the mean and dividing by the standard deviation (mean 0 and standard deviation 1).

The difference (95% confidence intervals [CI]) in mtDNA-CN by presence of asthma and by asthma severity in the UKB was evaluated using linear regression models adjusted for age, age$^2$, sex, center, log-transformed neutrophil and eosinophil counts, and platelet counts:

$$Adjusted\ mtDNA_{CN} = \beta_0 + \beta_1(asthma) + \beta_2(age) + \beta_3(age^2) + \beta_4(sex) + \beta_5 I(center) + \beta_6(\log neutrophil) + \beta_7(\log eosinophil) + \beta_8(platelet).$$

The relationship between mtDNA-CN and age was examined using the following linear models:

$$Unadjusted\ mtDNA_{CN} = \beta_0 + \beta_1(age) + \beta_2(asthma) + \beta_3(age \times asthma);$$

$$Unadjusted\ mtDNA\_CN = \beta_0 + \beta_1(age) + \beta_2(asthma) + \beta_3(age \times asthma) + \beta_4(sex) + \beta_5 I(center) + \beta_6(\log neutrophil) + \beta_7(\log eosinophil) + \beta_8(platelet).$$

Another two models were fitted by replacing asthma presence (no / yes) with asthma severity (non-asthma / mild asthma / moderate-severe asthma) in the above two models.

For SARP data, the adjusted mtDNA-CN was derived as the residual from the following regression model:

$$Unadjusted\ mtDNA_{CN} = cohort + age + age^2 + age * cohort + I(age^2) * cohort + sex + race + WBC + neutrophil.$$

The adjusted mtDNA CN was also standardized as the analyses for the UKB data.

The difference (95% confidence intervals [CI]) in mtDNA-CN by asthma severity was evaluated using linear regression models adjusted for the same set of covariates in the above model for adjusted CN.

The relationship between mtDNA-CN and age was examined using the following linear models:

$$Unadjusted\ mtDNA_{CN} = \beta_0 + \beta_1(age) + \beta_2(asthma\ severity) + \beta_3(age \times asthma\ severity);$$

$$Unadjusted\ mtDNA\_CN = \beta_0 + \beta_1(age) + \beta_2(asthma\ severity) + \beta_3(age \times asthma\ severity) + \beta_4(sex) + \beta_5 race + \beta_6 neutrophil + \beta_7 WBC.$$

**Table E3.** Features of asthmatics and healthy controls in SARP

| **Characteristics** | **Healthy Controls (*n* = 259)** | **Asthma (*n* = 1283)** | ***P* Value*** |
|---|---|---|---|
| Age — yr | 32 ± 11 | 42 ± 14 | <.0001 |
| Female — % | 63 | 66 | 0.3 |
| Ancestry — % | | | |
| European ancestry | 73 | 65 | 0.01 |
| African ancestry | 17 | 26 | 0.0009 |
| Body mass index — $kg/m^2$ | 26.5 ± 6.6 | 31.0 ± 8.4 | <.0001 |
| Blood pressure — mm Hg | | | |
| Systolic | 119 ± 14 | 125 ± 15 | <.0001 |
| Diastolic | 72 ± 9 | 76 ± 10 | <.0001 |
| Hemoglobin — g/dL | 14.2 ± 2.7 | 14.3 ± 5.2 | 0.7 |
| Hematocrit — % | 41.1 ± 5.6 | 41.3 ± 4.6 | 0.5 |
| Lung functions | | | |
| Forced expiratory volume in 1 second — % predicted | 98 ± 10 | 74 ± 21 | <.0001 |
| Forced vital capacity —% predicted | 99 ± 11 | 86 ± 18 | <.0001 |
| $FEV_1$/FVC | 0.83 ± 0.07 | 0.70 ± 0.11 | <.0001 |
| White blood cells — $x10^9$/liter | 6.1 ± 1.7 | 7.3 ± 2.5 | <.0001 |
| Neutrophils | 3.7 ± 3.0 | 4.4 ± 2.2 | 0.001 |
| Eosinophils | 0.13 ± 0.11 | 0.27 ± 0.24 | <.0001 |
| IgE — IU/ml | 73 ± 155 | 334 ± 700 | <.0001 |
| Glutathione peroxidase — U/ml | 0.087 ± 0.020 | 0.087 ± 0.023 | 0.9 |
| Superoxide dismutase — U/ml | 22.3 ± 10.8 | 19.1 ± 10.7 | 0.003 |
| Fractional exhaled nitric oxide — ppb | 18 ± 14 | 36 ± 35 | <.0001 |

Mean ± SD; **P* value for asthmatics *vs.* healthy controls.

**Table E4.** Features of study participants in quantile (Q)4 and Q1 of adjusted mtDNA-CN in SARP

| Characteristics | Q1 (*n* = 309) | Q4 (*n* = 310) | *P* Value* |
|---|---|---|---|
| Severe asthmatics — % | 43.0 | 42.2 | 0.8 |
| Age — yr | 39 ± 14 | 40 ± 14 | 0.5 |
| Female — % | 66 | 67 | 0.6 |
| Ancestry — % | | | |
| European ancestry | 67 | 68 | 0.7 |
| African ancestry | 28 | 26 | 0.6 |
| Body mass index — kg/$m^2$ | 29.8 ± 8.0 | 31.7 ± 8.6 | 0.005 |
| Blood pressure — mm Hg | | | |
| Systolic | 125 ± 15 | 124 ± 14 | 0.4 |
| Diastolic | 76 ± 11 | 76 ± 10 | 0.4 |
| Lung functions | | | |
| Forced expiratory volume in 1 second — % predicted | 74 ± 22 | 74 ± 20 | 0.9 |
| Forced vital capacity —% predicted | 87 ± 20 | 85 ± 17 | 0.3 |
| $FEV_1$/FVC | 0.70 ± 0.11 | 0.71 ± 0.12 | 0.3 |
| Asthma control test† | 17.4 ± 5.2 | 16.8 ± 4.9 | 0.3 |
| White blood cells — x$10^9$/liter | 7.1 ± 2.5 | 7.5 ± 3.0 | 0.08 |
| Neutrophils | 4.3 ± 2.2 | 4.6 ± 2.7 | 0.11 |
| Eosinophils | 0.26 ± 0.24 | 0.25 ± 0.20 | 0.5 |
| IgE — IU/ml | 360 ± 677 | 336 ± 765 | 0.6 |
| mtDNA-CN | 35.8 ± 18.9 | 178.4 ± 70.7 | <0.0001 |
| Unadjusted mtDNA-CN residuals | -0.747 ± 0.278 | 1.350 ± 1.040 | <0.0001 |
| Adjusted mtDNA-CN residuals | -0.947 ± 0.312 | 1.347 ± 1.024 | <0.0001 |

Mean ± SD; **P* value for Q4 *vs.* Q1 of adjusted mtDNA-CN. †Scores of 19 or higher are well controlled.

**Table E5.** Cutoff values for mtDNA-CN and other biomarkers with the exacerbation risk in asthmatics in SARP

| Variables | Exacerbations 1 – 2 *vs.* 0 | | | | Exacerbations ≥ 3 *vs.* 0 | | | | | | | | | |
|---|---|---|---|---|---|---|---|---|---|---|---|---|---|---|
| | OR1 | LR1 | UR1 | *P*1 | OR2 | LR2 | UR2 | *P*2 | Area under the ROC Curve | Optimal cutoff | OR3 | LR3 | UR3 | *P3* |
| Eosinophils— x10$^9$/liter | 1.093 | 0.872 | 1.369 | 0.441 | 1.447 | 1.144 | 1.832 | 0.002 | 0.602 | 0.338 | 3.073 | 1.730 | 5.459 | 0.000 |
| IgE — IU/ml | 0.883 | 0.699 | 1.114 | 0.294 | 0.781 | 0.538 | 1.134 | 0.194 | 0.600 | 45.000 | 0.420 | 0.234 | 0.757 | 0.004 |
| Superoxide dismutase — U/ml | 0.971 | 0.785 | 1.201 | 0.787 | 0.909 | 0.678 | 1.218 | 0.521 | 0.499 | 25.694 | 1.506 | 0.826 | 2.746 | 0.181 |
| Fractional exhaled nitric oxide — ppb | 1.137 | 0.905 | 1.429 | 0.269 | 1.387 | 1.090 | 1.764 | 0.008 | 0.589 | 27.500 | 2.134 | 1.213 | 3.755 | 0.009 |
| mtDNA-CN | 0.988 | 0.808 | 1.209 | 0.909 | 0.456 | 0.239 | 0.869 | 0.017 | 0.614 | 55.754 | 0.333 | 0.173 | 0.642 | 0.001 |
| Adjusted mtDNA-CN | 0.997 | 0.813 | 1.222 | 0.980 | 0.442 | 0.224 | 0.875 | 0.019 | 0.615 | -0.033 | 0.352 | 0.164 | 0.753 | 0.007 |

OR, odds ratio for exacerbation risk of asthma. LR and UR for upper and lower values of 95% confidence interval (CI). *P* value for the probability.

**Table E6.** Features of study participants who are divided by above and below mtDNA-CN cutpoint (-0.033) in SARP

| **Characteristics** | **Below (*n* = 794)** | **Above (*n* = 445)** | ***P* Value*** |
|---|---|---|---|
| Severe asthmatics — % | 46.1 | 43.3 | 0.3 |
| Age — yr | 42 ± 14 | 40 ± 15 | 0.04 |
| Female — % | 65 | 67 | 0.5 |
| Ancestry — % | | | |
| European ancestry | 65 | 66 | 0.7 |
| African ancestry | 27 | 26 | 0.8 |
| Body mass index — $kg/m^2$ | 30.8 ± 8.4 | 31.6 ± 8.4 | 0.11 |
| Blood pressure — mm Hg | | | |
| Systolic | 125 ± 15 | 123 ± 14 | 0.2 |
| Diastolic | 77 ± 10 | 76 ± 9 | 0.5 |
| Lung functions | | | |
| Forced expiratory volume in 1 second — % predicted | 74 ± 21 | 74 ± 20 | 0.6 |
| Forced vital capacity —% predicted | 86 ± 19 | 86 ± 18 | 0.9 |
| $FEV_1$/FVC | 0.69 ± 0.12 | 0.70 ± 0.12 | 0.18 |
| Asthma control test† | 17.4 ± 4.9 | 16.8 ± 4.9 | 0.13 |
| White blood cells — $x10^9$/liter | 7.2 ± 2.4 | 7.5 ± 2.8 | 0.09 |
| Neutrophils | 4.3 ± 2.0 | 4.5 ± 2.4 | 0.3 |
| Eosinophils | 0.28 ± 0.25 | 0.26 ± 0.22 | 0.2 |
| IgE — IU/ml | 336 ± 713 | 331 ± 682 | 0.8 |
| mtDNA-CN | 51.2 ± 23.4 | 149.8 ± 74.2 | <0.0001 |
| Unadjusted mtDNA-CN residuals | -0.520 ± 0.359 | 0.930± 1.092 | <0.0001 |
| Adjusted mtDNA-CN residuals | -0.534 ± 0.395 | 0.954± 1.042 | <0.0001 |

Mean ± SD; **P* value for Above *vs.* Below adjusted mtDNA-CN cutoff value of -0.033. †Scores of 19 or higher are well controlled.

### Supplementary Figures

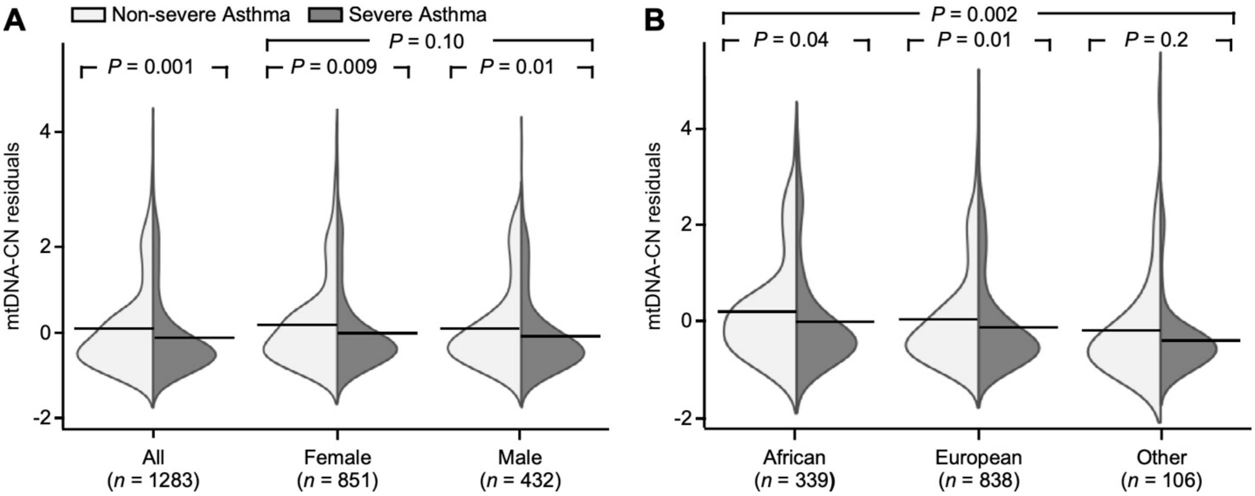


**Figure E1.** Severe asthmatics have significantly lower mtDNA-CN than non-severe asthmatics in the Severe Asthma Research Program (SARP). (*A*) Lower mtDNA-CN in severe asthmatics by sex. Lower p-values from comparison of non-severe vs severe asthma overall, in females, and in males. Upper p-value from comparison of all females vs all males. (*B*) Lower mtDNA-CN in severe asthmatics by ancestry. Lower p-values from comparison of non-severe vs severe asthma in individuals of African ancestry, individuals of European ancestry, and individuals of other ancestries. Upper p-value from comparison of ancestry.

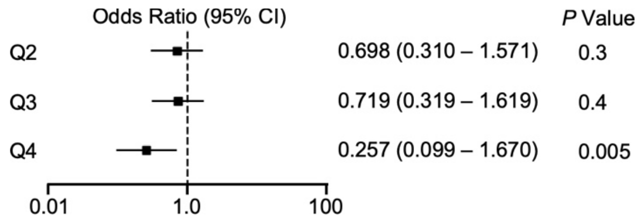


**Figure E2.** Odds ratio of risk of three or more exacerbations by quantile of adjusted mtDNA-CN in asthmatics in SARP.

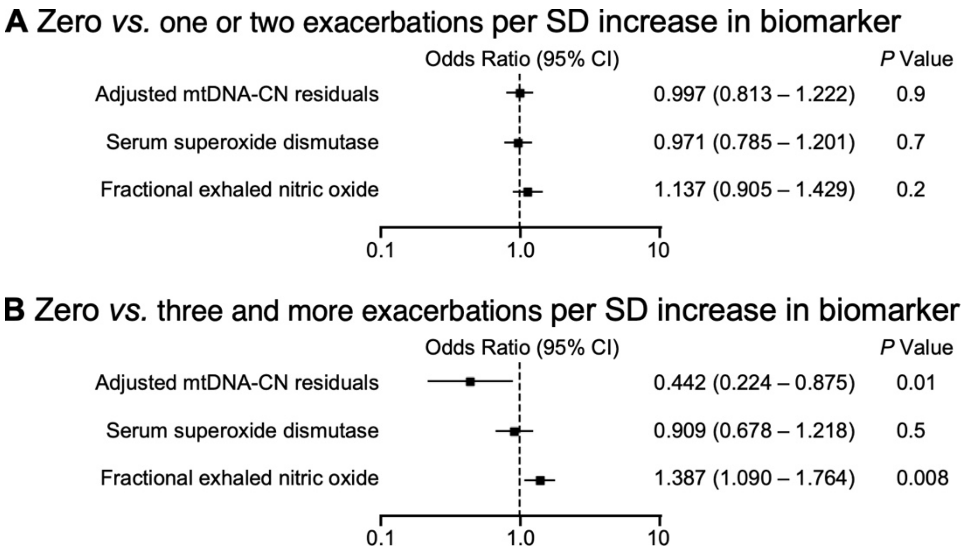


**Figure E3.** Associations of adjusted mtDNA-CN and other biomarkers with the exacerbation risk in asthmatics in SARP. (*A*) Zero exacerbations *vs.* one or two exacerbations per SD increase in biomarker. (*B*) Zero exacerbations *vs.* three and more exacerbations per SD increase in biomarker.
